# Longitudinal SARS-CoV-2 testing is punctuated by intermittent positivity and variable rates of cycle-threshold decline

**DOI:** 10.1101/2021.10.01.21264373

**Authors:** Shawn E. Hawken, Subhashini A. Sellers, Jason R. Smedberg, Jeremy D. Ward, Herbert C. Whinna, William Fischer, Melissa B. Miller

## Abstract

The COVID-19 pandemic is complicated by cases of vaccine-breakthrough, re-infection, and widespread transmission of variants of concern (VOC). Consequently, the need to interpret longitudinal positive SARS-CoV-2 (SCV-2) tests is crucial in guiding clinical decisions regarding infection control precautions and treatment. Although quantitative tests are not routinely used diagnostically, standard diagnostic RT-PCR tests yield Ct values that are inversely correlated with RNA quantity. In this study, we performed a retrospective review of 72,217 SCV-2 PCR positive tests and identified 264 patients with longitudinal positivity prior to vaccination and VOC circulation. Patients with longitudinal positivity fell into two categories: short-term (207, 78%) or prolonged (57, 22%) positivity, defined as <= 28 (range 1-28, median 16) days and >28 (range 29-152, median 41) days, respectively. In general, Ct values declined over time in both groups; however, 11 short-term positive patients had greater amounts of RNA detected at their terminal test compared to the first positive, and 5 patients had RNA detected at Ct < 35 at least 40 days after initial infection. Oscillating positive and negative results occurred in both groups, although oscillation was seen three times more frequently in prolonged-positive patients. Patients with prolonged positivity had diverse clinical characteristics but were often critically ill and were discharged to high-level care or deceased (22%). Overall, this study demonstrates that caution must be emphasized when interpreting Ct values as a proxy for infectivity, predictor of severity, or a guide for patient care decisions in the absence of additional clinical context.

## Background

The global burden of morbidity and mortality caused by SARS-CoV-2 (SCV-2) is concentrated among vulnerable patients with comorbidities (1). While most patients test negative after 14 days, cases where patients remain RNA positive months later have been observed (2–5). The recent emergence of vaccine-breakthrough cases, re-infections, and widespread transmission of variants of concern (VOC), such as World Health Organization lineage Delta, which appears to rise to higher viral titers, underscore the clinical imperative to interpret positive SCV-2 results from the same patient over time. Unfortunately, current gold-standard RT-PCR clinical diagnostic tests are not quantitative (6, 7). While RT-PCRs yield Ct values that are inversely correlated with the amount of RNA present in a sample, they are influenced by patient, specimen, and diagnostic test characteristics (8, 9).

The debate over if and how to use Ct values clinically is underscored by recent statements from professional societies including the joint statement from the Infectious Diseases Society of America (IDSA) and the Association for Molecular Pathology (AMP) and a statement from the College of American Pathologist (CAP), in response to growing interest in using Ct values as surrogate measure of a patients viral load (8, 9). Proponents of using Ct values to guide clinical decisions infer Ct values as a snapshot of viral burden in a patient’s sample, and there is interest in clinical evaluation of Ct trends to discern the clinical and epidemiological relevance of a positive test (9, 10). Unfortunately, the observation that a subset of patients will test positive, typically with a higher Ct value several months after initial infection are a challenge clinically, since it is often unclear if these patients are still infectious and what distinguishes these patients from the majority of SCV-2 patients who instead convert to a negative test weeks after infection (3–5). Here we sought to describe the natural history of SCV-2 test results, including Ct values, among patients who tested positive multiple times at our institution in a time period prior to vaccine roll-out and VOC circulation. We describe the duration of positivity and clinical characteristics of the population of patients with prolonged SCV-2 positivity. This investigation serves to highlight challenges in using Ct values to guide clinical decisions.

## Methods

### Study design and participants

We performed a descriptive retrospective review of all patients who underwent testing for SCV-2 RNA at the University of North Carolina Medical Center Clinical Microbiology Laboratory between March 17, 2020 and October 15, 2020. This study was approved by the Institutional Review Board at the University of North Carolina at Chapel Hill (IRB# 20-2448).

### SARS-CoV-2 RNA testing

During the study timeframe, four platforms were used by our laboratory for the detection of SCV-2 RNA. Three of these assays, the Abbott Alinity m SARS-CoV-2 assay, Abbott RealTime SARS-CoV-2 assay (Abbott Molecular Inc, Green Oaks, IL), and Cepheid Xpert® Xpress SARS-CoV-2 (Cepheid, Sunnyvale, CA) had accessible cycle threshold data that was exported from the instruments. Ct values were not available from an Emergency use authorization (EUA) laboratory developed test that was used on a minority of specimens. To adjust for cycle threshold reporting differences between instruments, 10 cycles were added to Ct values from the Abbott M2000 assay to account for the 10 hidden cycles in the analysis software (Abbott Molecular Inc, personal communication).

### Clinical chart review and data abstraction

Basic demographic information and SCV-2 test results (positive and negative tests, specimen type, testing platform, Ct values) were abstracted from electronic medical records (EMR) and the laboratory information system for all patients tested for SCV-2 RNA during the study period. We queried these reports to identify patients with multiple positive tests and examined the timing of positive tests to identify patients who had evidence of long-term positivity of SCV-2 RNA. Patients who demonstrated positivity later than the 3^rd^ quartile of all patients with multiple positive tests were considered “long-term” positive patients and were investigated further by chart review. Manual chart review of the EMR was performed to abstract patient age, gender, body mass index (BMI), hospital admission dates, and discharge disposition. Medication exposure for long-term positive patients was collected via EMR reports.

SCV-2 testing data were evaluated in relation to positive and negative testing dates to describe the characteristics of the patient population with prolonged positivity and to summarize patterns of clinical exposures surrounding dates where patients either tested negative or positive for SCV-2.

### Statistical analysis

All descriptive statistics and data analysis were performed using R version 3.6.1 (11).

## Results

### Summary of SARS-CoV2 testing during study

Between March 16 and October 15, 2020 our hospital performed 72,217 RT-PCR tests for SCV-2 including 4,609 positive and 67,608 negative tests across six different specimen types including 67,612 (93%) nasopharyngeal swabs. RT-PCR tests were performed across four platforms: 18,057 EUA LDT, 7,014 Cepheid Xpert, 23,983 Abbott Alinity m, and 23,163 Abbott RealTime (m2000). Among the 58,348 patients tested, 9,682 (16.6%) were tested multiple times and 264 (0.45%) patients had at more than one positive test (**Supplemental Figure 1**). Among 264 patients with multiple positive tests, the median interval between first and last positive test was 16 days (range 1-152) with 118 (44.7%) of patients first and last testing positive within a single 14-day incubation period. We took advantage of a natural breakpoint in the distribution of duration of positivity to define prolonged positivity as >3^rd^ quartile (28 days) duration between first and last positive tests (**Figure 1**). Prolonged SCV-2 positivity was observed in 57 patients who remained positive for 29-152 (median 41) days.

**Figure 1.**
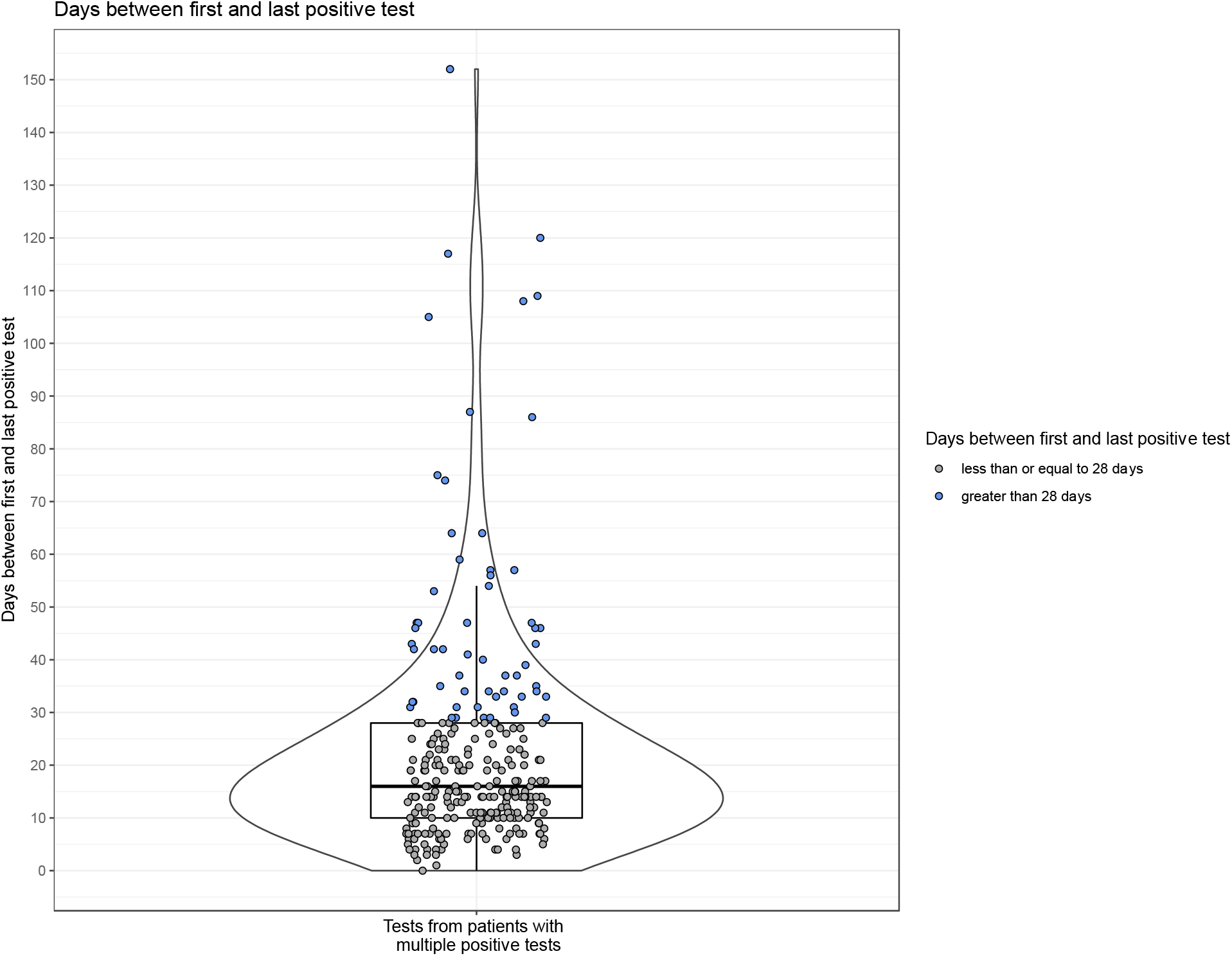
Summary of time between first and last positive test for patients with multiple positive tests. Y-axis indicates days between first and last positive test, individual dots indicate individual patients. Blue indicates prolonged positive patient defined at natural breakpoint of > 3^rd^ quartile duration, grey indicates short-term positive patient. Violin and box plot indicate overall distribution of days between first and last positive test.

### Diagnostic testing characteristics of patients with multiple SCV-2 positive tests

Among the 207 patients with multiple positive tests who did not have prolonged positivity, 17 (8.2%) had patterns of intermittent positivity with at least one negative test (range 0-3 negative tests) between their first and last positive test. This pattern was observed over three-times more frequently among the 57 prolonged positive patients among whom 16 (28%) had intermittent positivity (p<0.001).

While Ct values generally declined over time, variable rates of decline in Ct values were observed in individual patients (**Figure 2 A-B, Supplemental Figure 2**). Among patients without prolonged positivity, 123 (59%) had Ct values available from both the first and last (terminal) positive test. Among these patients, terminal positive tests were median 9.8 cycles higher compared to first positive tests, signaling lower amounts of RNA detected; however, 11 patients had higher amounts of RNA detected at their last positive test compared to first positive test which occurred between 3 and 25 days later. Overall, terminal positive tests ranged from 22.5 cycles lower to 31.8 cycles higher, with terminal positives testing near the limit of detection (>35) for 21 (17%) patients. For prolonged positive patients’ terminal and initial test Ct data was available for 36 patients (63%). Terminal positive tests were a median of 14 (range 2-26) cycles greater than initial positive tests.

**Figure 2.**
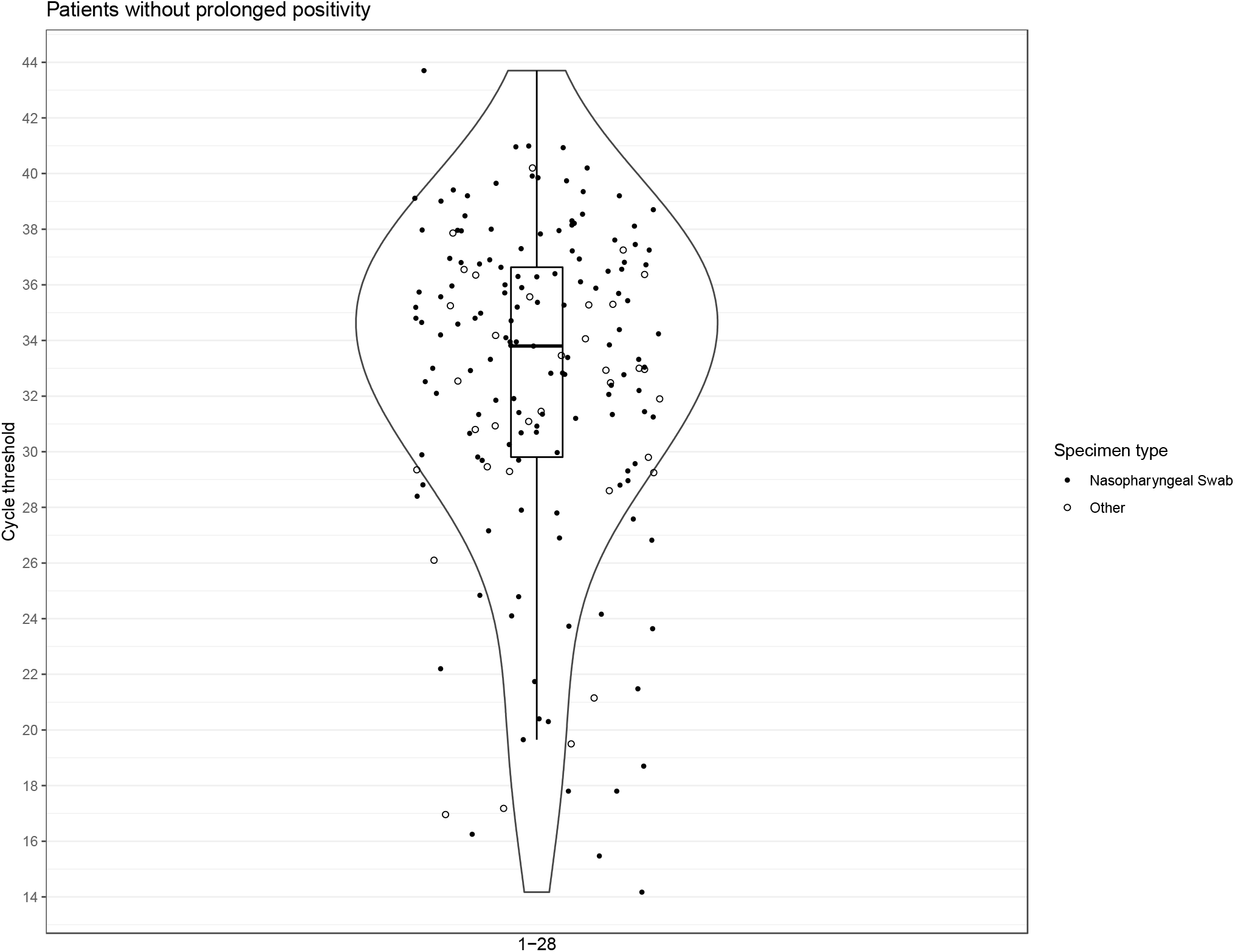
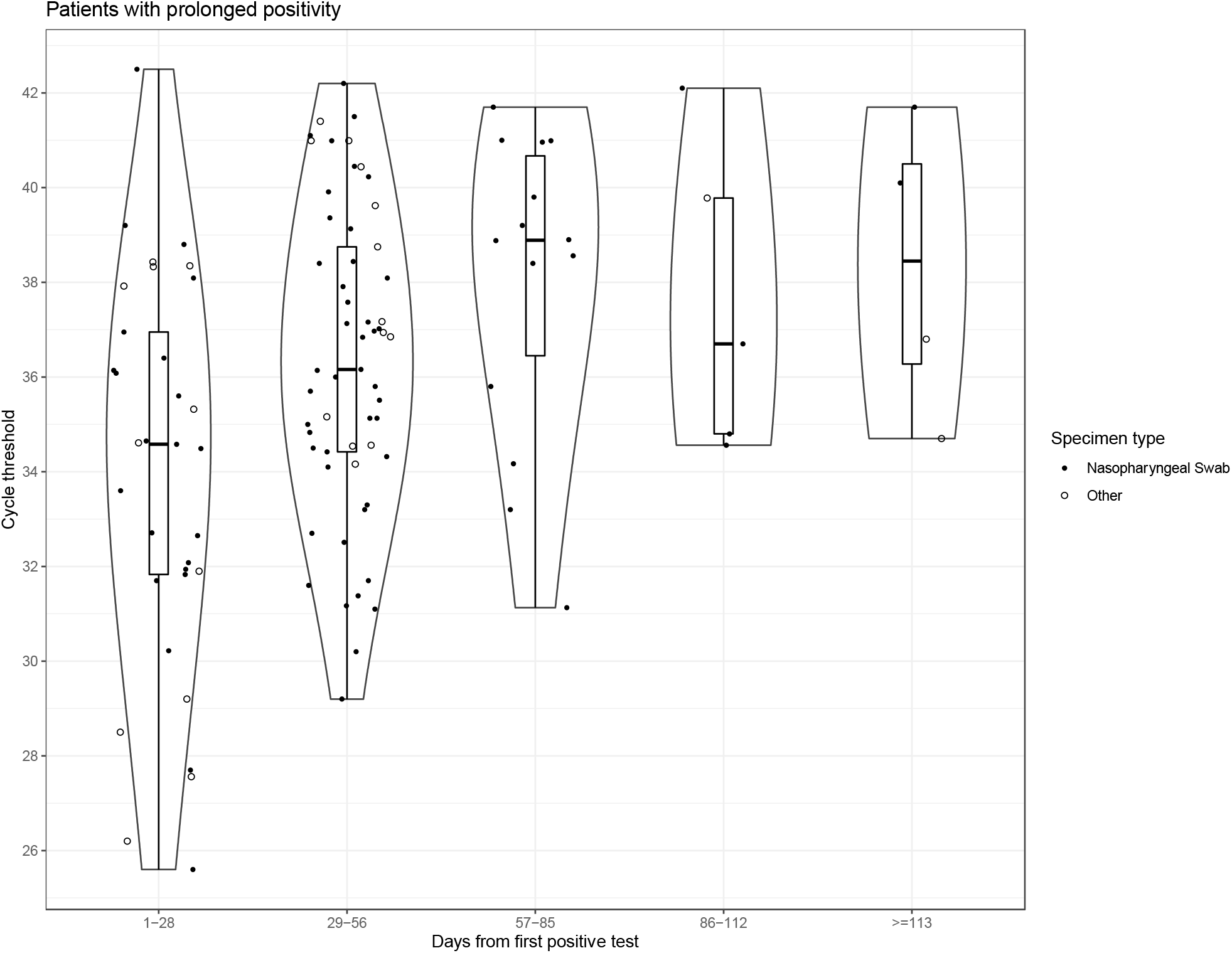
Distribution of Ct values over time for patients with and without prolonged positivity. X-axis indicates days between positive tests, y-axis indicates cycle-threshold of clinical tests. A. patients without prolonged positivity. B. patients with prolonged positivity. Symbols indicate nasopharyngeal swab or other specimen type. Violin and box plot indicate distribution of Ct values at timeframe specified on x-axis.

In contrast to patients without prolonged positivity, no prolonged positive patients had terminal test Ct values that were lower than their initial positive test (i.e., increase in amount of RNA detected). Consistent with continued viral RNA decline over time, 33 (91%) prolonged positive patients had terminal positive tests near the limit of detection (>35 Ct); however, by day 40 post initial positive test, 5 (13.8%) patients still had Ct values <35 and one patient tested positive with a Ct value <35 109 days after initial positive (**Figure 3**).

**Figure 3.**
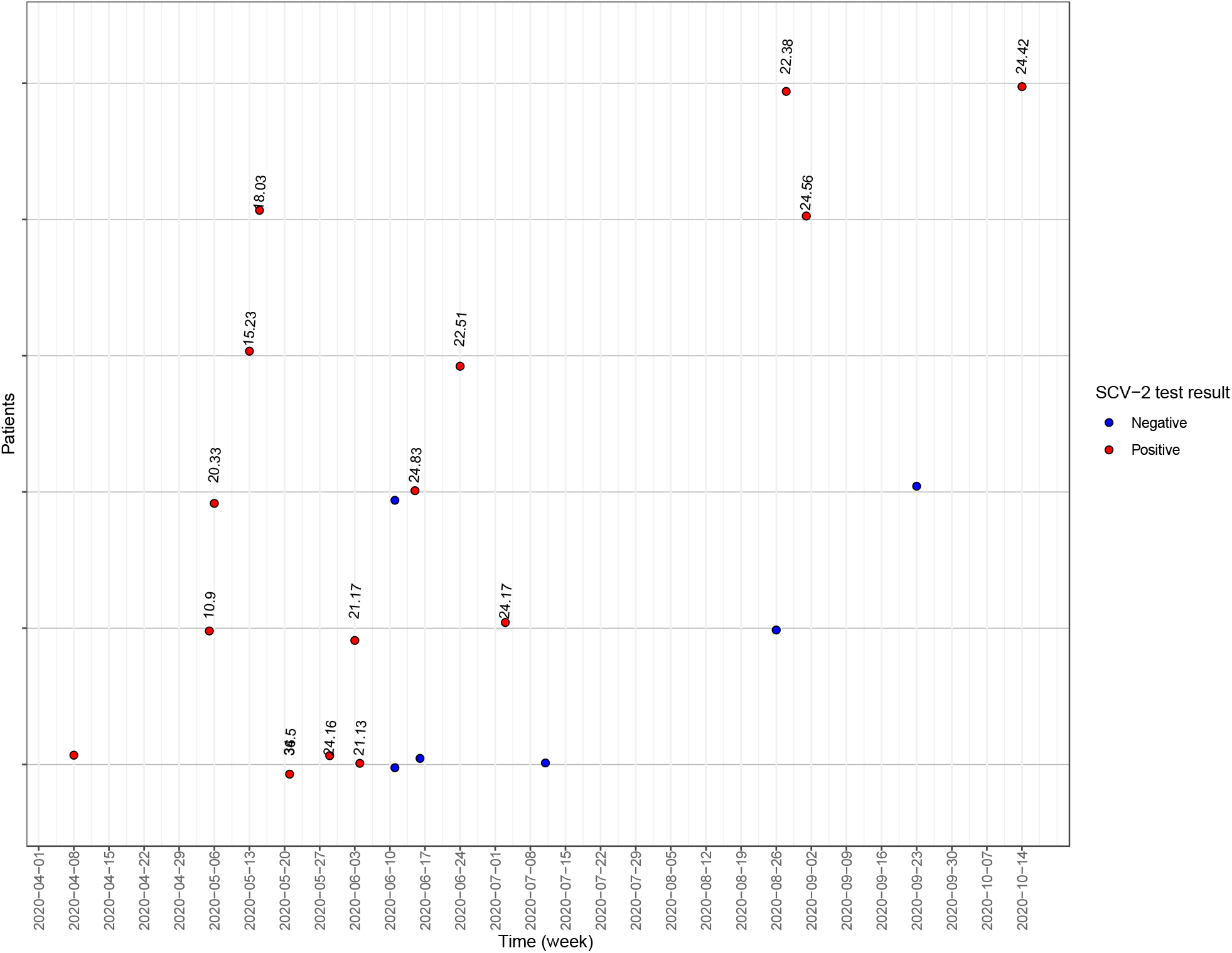
SCV-2 testing summary for patients with slow Ct value decline defined as Ct values <35 after 40 days post initial positive test. X-axis indicates time, y axis indicates 1 patient per grey line. Red and blue dots indicate positive and negative SCV-2 tests respectively. Table 1. Clinical characteristics of patients with prolonged SCV-2 positivity.

### Demographic and treatment characteristics of prolonged positive patients

Patients with prolonged positivity are a population where the examination of Ct values is an attractive method to help guide clinical decisions. Clinical characteristics of the 57 prolonged positive patients included: obesity 30%, hospitalization 53%, discharge to long-term care 35%, hospice or deceased 23%. In general, prolonged positive patients experienced a severe course of illness, with 33% treated with immune modulating drugs and 10% treated with remdesivir (**Table 1**).

**Table 1.** Clinical characteristics of patients with prolonged SCV-2 positivity.

|  | Overall<br>(N=57) |
| --- | --- |
| **Sex** |  |
| F | 31 (54 %) |
| M | 26 (46 %) |
| **BMI (kg/m^2)** |  |
| Mean (SD) | 31 (± 9.8) |
| Missing | 25 (43.9%) |
| **Age (years)** |  |
| Mean (SD) | 49 (± 20) |
| Missing | 1 (1.8%) |
| **Cycle threshold first positive** |  |
| Mean (SD) | 23 (± 6.0) |
| Missing | 13 (22.8%) |
| **Cycle threshold last positive** |  |
| Mean (SD) | 37 (± 3.2) |
| Missing | 11 (19.3%) |
| **Time between first and last positive (days)** |  |
| Mean (SD) | 51 (± 27) |
| **Admitted** |  |
| admitted | 30 (53 %) |
| never admitted | 27 (47 %) |
| **Discharge_disposition** |  |
| Assisted Living | 2 (4 %) |
| Deceased | 1 (2 %) |
| Home | 17 (30 %) |
| Hospice | 1 (2 %) |
| Never Admitted | 27 (47 %) |
| Rehab | 1 (2 %) |
| SNF | 6 (11 %) |
| Missing | 2 (3.5%) |

## Discussion

Here we describe the natural history of SCV-2 RNA testing among all patients who tested positive multiple times at our institution during a seven-month period, revealing complex patterns of intermittent positivity and variable decline of viral RNA detected via Ct values throughout different stages of infection. Although factors observed among prolonged positive patients were similar to those described in patients with severe COVID disease, 47% of prolonged positive patients were not ill enough to require hospitalization, underscoring that a large subset of healthier patients may experience prolonged RNA positivity (1, 12). Intriguingly, 11 patients without prolonged positivity demonstrated greater amounts of RNA detected at their terminal versus initial SCV-2 positive test, suggesting variable shedding even early in their infection (2). Additional factors such as timing of initial presentation and specimen quality, likely contribute to RNA detection or observed decline (2, 5, 13, 14). Although it was beyond the scope of this study to evaluate likelihood of re-infection, which would require longitudinal viral sequencing, the observation of decreased RNA detection over time in the majority of patients was consistent with a single infection course, since re-infection is currently thought to be rare, especially prior to widespread VOC circulation (18).

The observation of intermittent positivity in 8% of patients with multiple SCV-2 positive tests during a time period prior to sustained VOC transmission suggests sample quality may play a major role in Ct results (13). Interestingly, prolonged positive patients were over three times more likely to have intermittent positivity. These intermittent negatives may be explained by variable shedding dynamics, therapeutic interventions, sample quality, among other variables, which will need to be investigated in future studies in order to better understand this phenomenon and how it pertains to infectivity and the clinical course of these patients.

As the pandemic progresses in time, and VOC capable of high-titer and vaccine breakthrough infections such as Delta gain in prevalence, longitudinal testing information with variability in Ct values will become available for a greater number of patients. With the added complexity of novel variant circulation, it will become even more crucial to keep the possibility of variable shedding in mind at any stage of infection and exercise caution when interpreting Ct values as proxy measures for infectivity and severity. Development and deployment of diagnostic tests that can discriminate between prolonged shedding and reinfection and provide insight into infectiousness is urgently needed for the next stage of the SCV-2 pandemic.

## Data Availability

Not applicable

## Acknowledgments

We would like to thank the staff of the Clinical Microbiology and Molecular Microbiology Laboratories at UNC Medical Center for their tireless efforts during the pandemic and their role in performing the clinical SCV-2 tests described in this study.

## Supplemental Information

**Figure S1.**
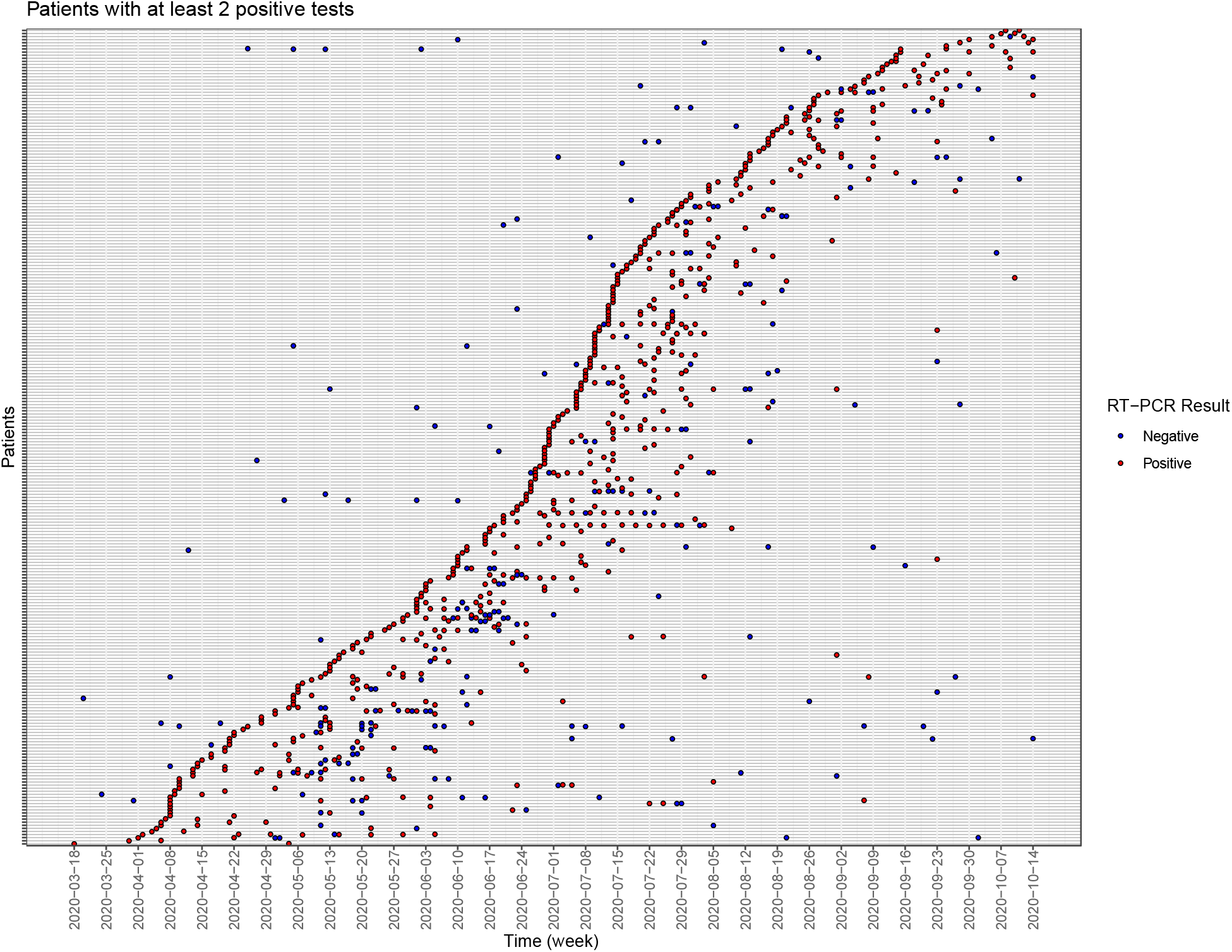
Timeline of SCV-2 testing throughout study for patients with at least 2 positive SCV-2 tests. X-axis indicates time, y axis indicates 1 patient per grey line. Red and blue dots indicate positive and negative SCV-2 tests respectively.

**Figure S2.**
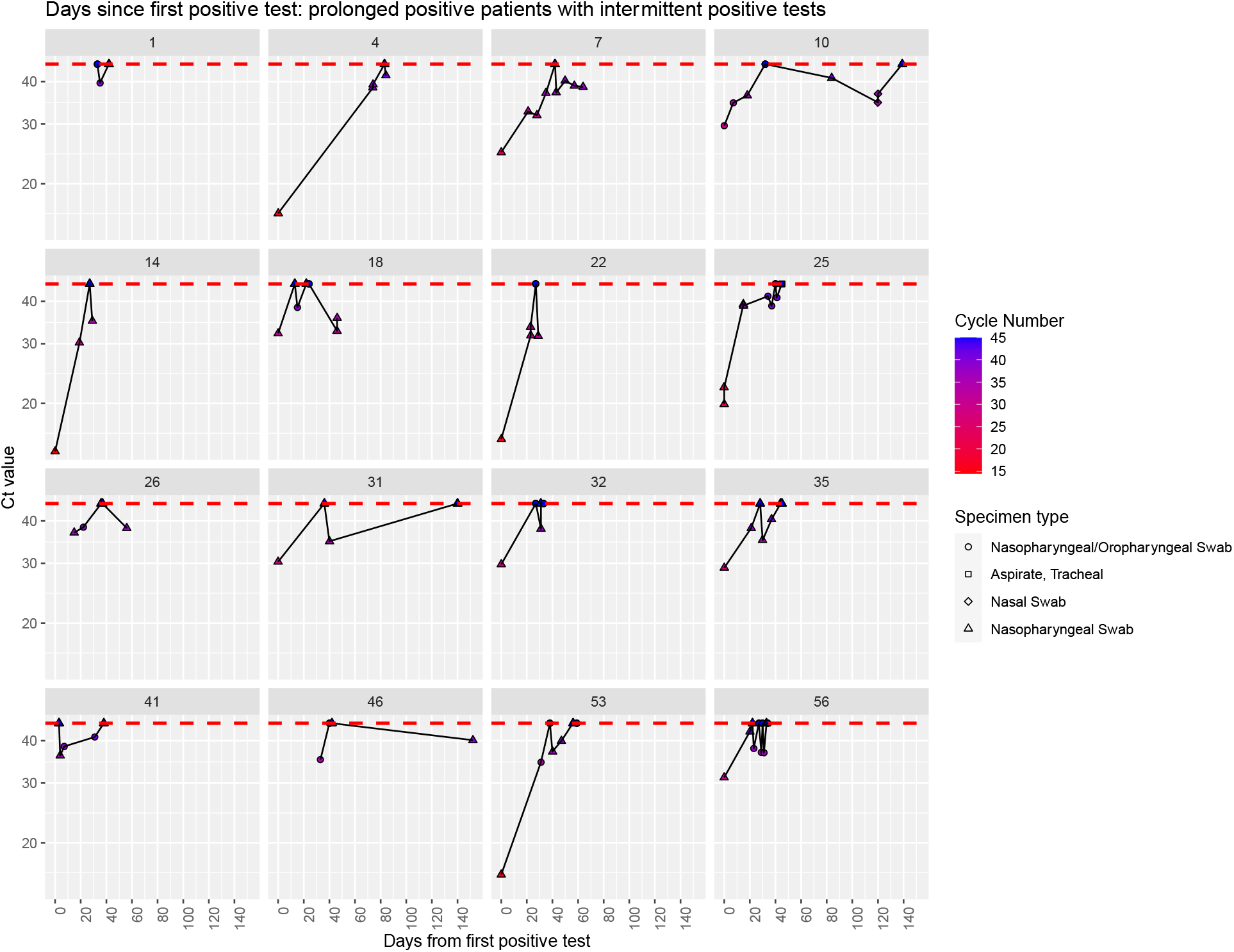
Timeline of Ct values for prolonged positive patients with intermittent positive and negative tests. Y axis indicates Ct value, X axis indicates days from first positive test. Patients are represented by number in grey box at top of each plot. Red line indicates negative test. Tests are only plotted if negative or positive and a Ct value was available.

